# Rapid estimation of excess mortality in times of COVID-19 in Portugal Beyond reported deaths

**DOI:** 10.1101/2020.05.14.20100909

**Authors:** André Vieira, Vasco Peixoto Ricoca, Pedro Aguiar, Alexandre Abrantes

## Abstract

**Background:** One month after the first COVID-19 infection was recorded, Portugal counted 18 051 cases and 599 deaths from COVID-19. To understand the overall impact on mortality of the pandemic of COVID-19, we estimated the excess mortality registered in Portugal during the first month of the epidemic, from March 16 until April 14 using two different methods.

**Methods:** We compared the observed and expected daily deaths (historical average number from daily death registrations in the past 10 years) and used 2 standard deviations confidence limit for all-cause mortality by age and specific mortality cause, considering the last 6 years. An adapted ARIMA model was also tested to validate the estimated number of all-cause deaths during the study period.

**Results:** Between March 16 and April 14, there was an excess of 1,255 all-cause deaths, 14% more than expected. The number of daily deaths often surpassed the 2 standard deviations confidence limit. The excess mortality occurred mostly in people aged 75+. Forty-nine percent (49%) of the estimated excess deaths were registered as due to COVID-19, The other 51% registered as other natural causes.

**Conclusion:** Even though Portugal took early containment measures against COVID-19, and the population complied massively with those measures, there was significant excess mortality during the first month of the pandemic, mostly among people aged 75+. Only half of the excess mortality was registered as directly due do COVID-19.

## 1 Background

The COVID-19 pandemic is expected to cause excess mortality (EM) throughout 2020, both directly, due to deaths among those infected, and indirectly, due to patients’ not seeking health care for fear of becoming infected by COVID-19, or due to the inability of healthcare system to provide effective services to patients other than those with COVID-19. The economic and social impact of the pandemic could also contribute to long-term EM.^1^

According to Johns Hopkins University, by April 15, 2020, the COVID-19 virus had infected nearly 2 million people and killed more than 128,000 people worldwide.^2^ EuroMOMO reports a significant increase in the number of deaths caused by COVID-19 in several countries such as Spain and Italy.^3^ Other analyzes, for countries like England, France, Switzerland and the Netherlands already presented considerable excess mortality as of 12 April.^4^ These values would probably have been much higher if no action had been taken. However, making comparisons between different methods can be challenging, since the methods used and periods considered vary considerably, from study to study.

On March 16, 2020, the first death due to COVID-19 was registered in Portugal, 14 days after the first confirmed case of infection. In March 2020, 187 people died of COVID-19, or 2.3% of the 8 521 confirmed patient cases, a cumulative incidence of around 80 cases per 100 000 inhabitants and a lethality rate of 2.3%. In Portugal, most deaths by COVID-19 reported so far have occurred in individuals with underlying health conditions or at older ages^5^,

This article estimates excess mortality during the first month of recorded deaths by COVID-19.

## Method

Mortality data was extracted from the Portuguese Death Certificate Information System (SICO-eVM).^6^ Data on the number of COVID-19 reported deaths in Portugal were extracted from information in the DGS (Directorate-General of Health) daily Situation Reports.
^7^

For estimating total excess mortality (EM), we calculated the historical average and respective standard deviation (SD) of the number of daily all-cause mortality for the last 10 years, between January 1, and April 14. To estimate the excess mortality by age and cause (natural and external), we used only the last six years average, because the information was not available for earlier years.

We considered Relevant Excess Mortality (REM) when the daily observed values exceed the estimated average value of deaths for each day plus corresponding 2 SDs or if they exceed the limits of the 95% confidence interval in the ARIMA model. The REM calculation method has been used in similar studies.^8, 9^

To validate the EM calculated on the basis of historical daily deaths, from all causes, we compared it with the estimated results obtained with an ARIMA model (1,1,5) adjusted to the time series until April 14, 2020. We modeled a time series including the observed mortality from January 1, 2010 to March 15, 2020 and the historical average after that day from March 16, 2020 until 14 April 2020. This cut-off point (March 16) was the date of first reported COVID-19 death and the close inversion in mortality trend. We compared the observed and modeled expected mortality for the pandemic period and the respective 95% confidence intervals. The model was adjusted with the SPSS program, with a determination coefficient of R2 = 0.801, Ljung-Box test not statistically significant, Autocorrelation Function and Partial Autocorrelation Function not statistically significant in the residues, and estimates of the auto parameters - statistically significant regression and moving averages (p <0.01).^10^

## 2 Results

### All-cause excess mortality

Between March 16 and April 14, 2020, there was an excess of 1 255 deaths over the expected, based on average daily mortality during the previous 10 years, an excess mortality of 13.7%. The number of deaths observed exceeded the defined threshold of Relevant Excess Mortality in 14 days of the study period, totaling 242 REM.

**Figure 1.**
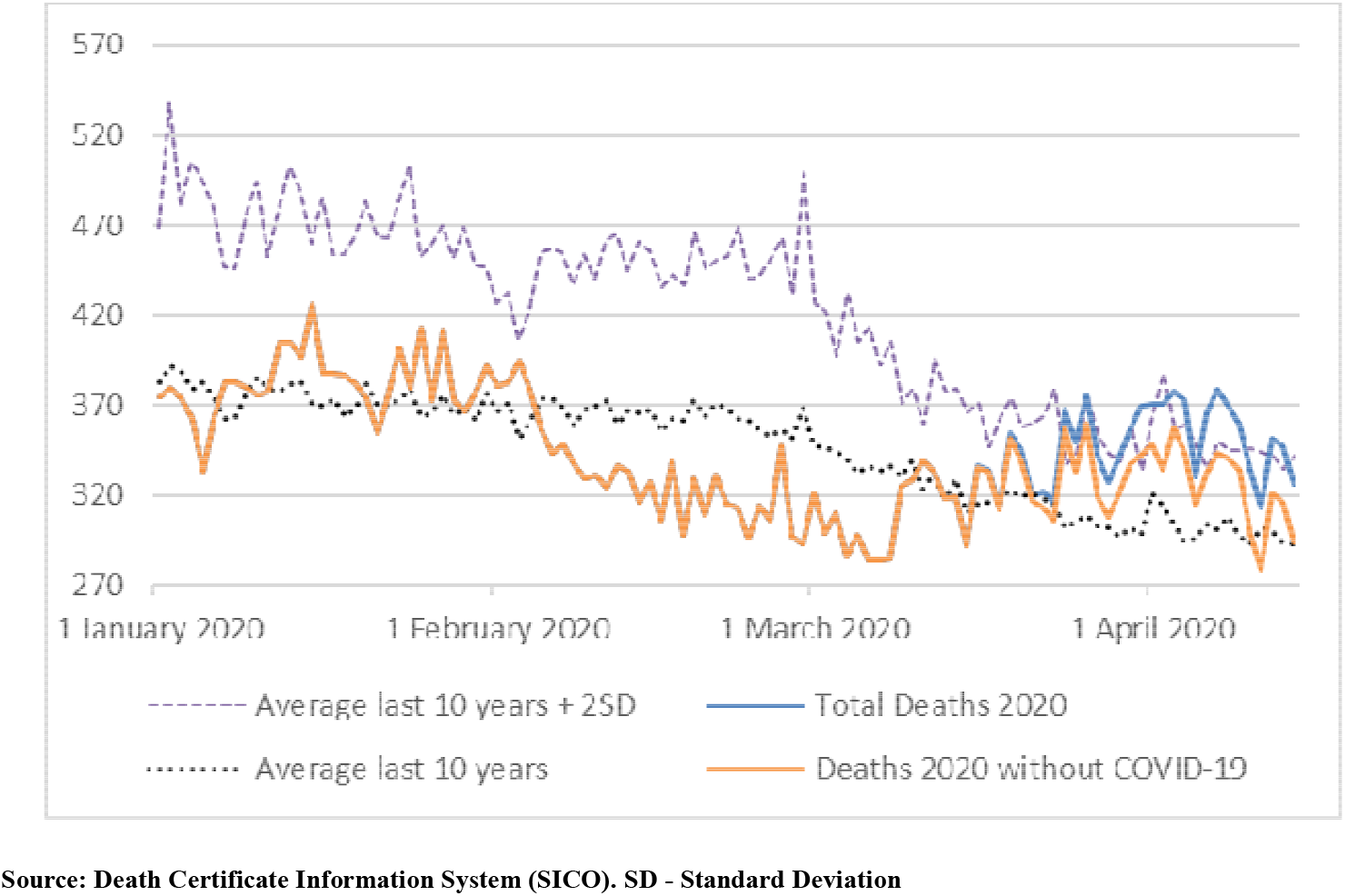
All-cause mortality between 1st January and 14th April 2020

During the 10 years’ time-series of mortality data, March and April showed declining mortality in relation to January and February. In 2020, there is an inversion of this negative trend as of March 11, with an EM above the average of the previous 10 years, exceeding the average threshold plus 2SD, as of March 24, 2020.

### Excess mortality by age

Almost all excess mortality (EM) was detected among people aged 75 years or more. Between March 1 and April 14, 2020, there was an EM of 1 030 deaths in people aged 75 +, compared to the daily averages of deaths in this age group, in the last 6 years. For this age group, the number of deaths observed exceeded the defined threshold of Relevant Excess Mortality in 11 days of the study period, totaling 201 REM.

In the same period, only 67 deaths were estimated above the historical average in the age group between 65 and 74 years old and EM was not identified in any other age group.

### Excess mortality by cause

The COVID-19 reported deaths are included in deaths due to natural causes. Between March 16 and April 14, there was an EM of 1 281 deaths from natural causes, based on deaths registered in the last 6 years. (Table 1 and Figure 2)

**Figure 2.**
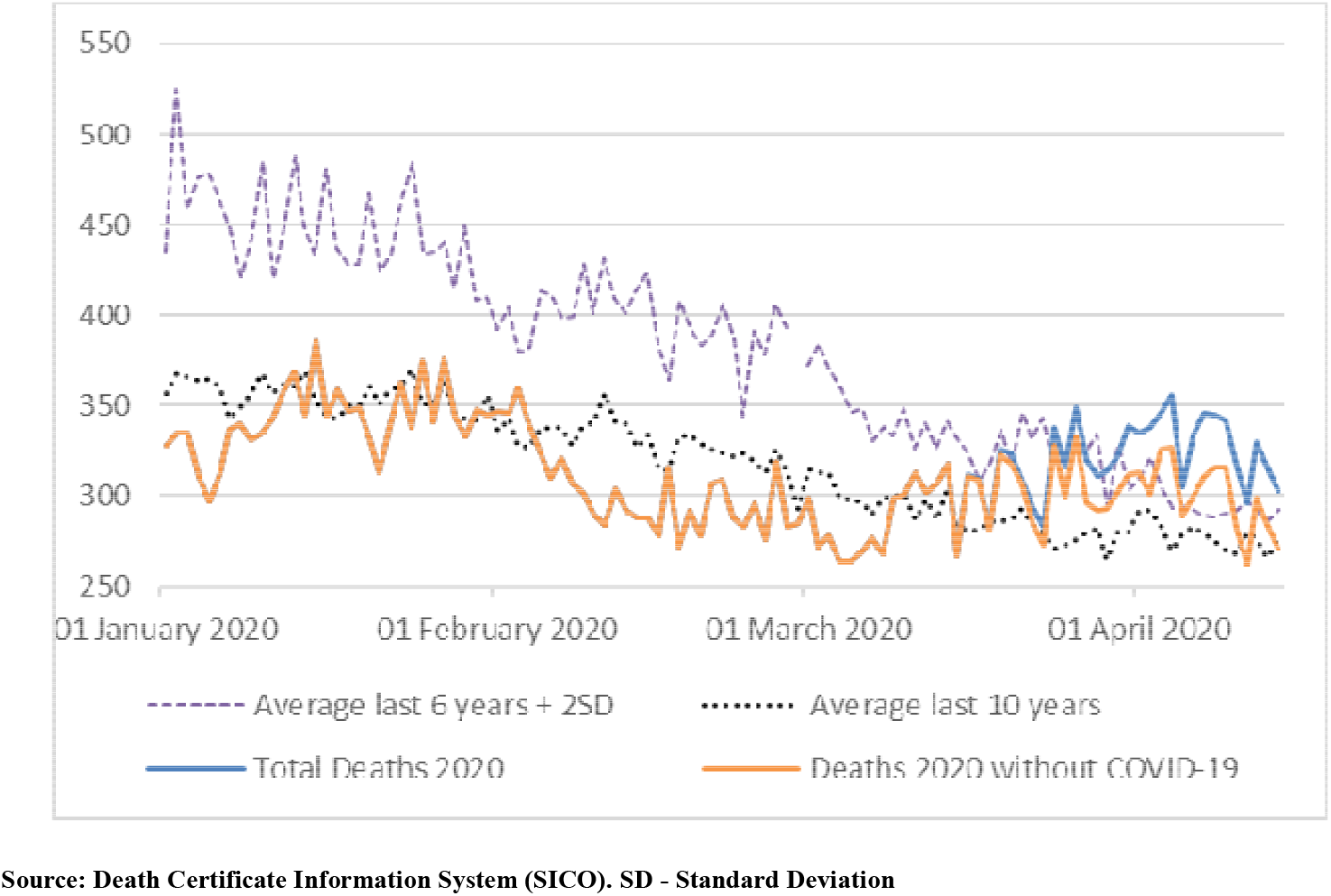
Mortality by natural cause between 1st January and 14th April 2020

The number of deaths due to natural causes exceeded the average +2SD limit for the last 6 years in 19 days, totaling 566 deaths in REM. Between March 16 and April 14, there was a 15.2% increase in deaths from natural causes and a 57% reduction in deaths from external causes compared to the averages observed in the previous 6 years.

### ARIMA modelling

Forty nine percent of excess deaths were caused by COVID-19 and another 51% were attributed to other natural causes.

Comparing the observed deaths with those modeled for March and April, we observed an EM from March 16, exceeding the upper limit of the confidence interval in the last days of March and the first days of April (Figure 3). There is also a tendency towards a reduction in mortality in April, as some effect of the confinement measures is already expected to have occurred. Between March 16 and April 14, there was an excess of 1214 deaths in relation to the deaths those estimated for a situation in which there was no epidemic. 10445 deaths were observed and the chronological model estimates that 9231 deaths would have been recorded, had there not been a pandemic. Excess mortality was similar in the two applied methods suggesting that results of this study are robust.

**Figure 3.**
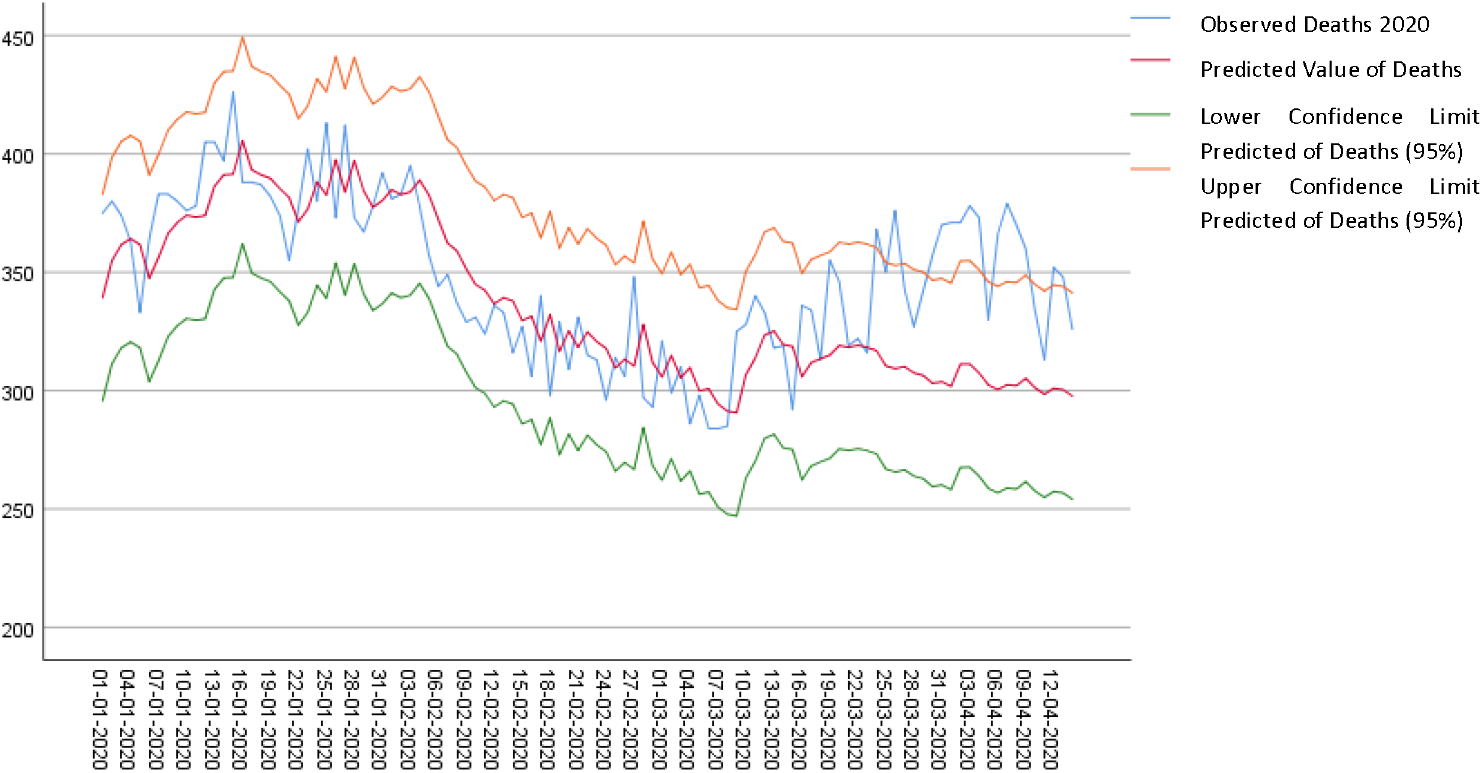
Evolution of mortality analyzed using the ARIMA model.

**Table 1.**
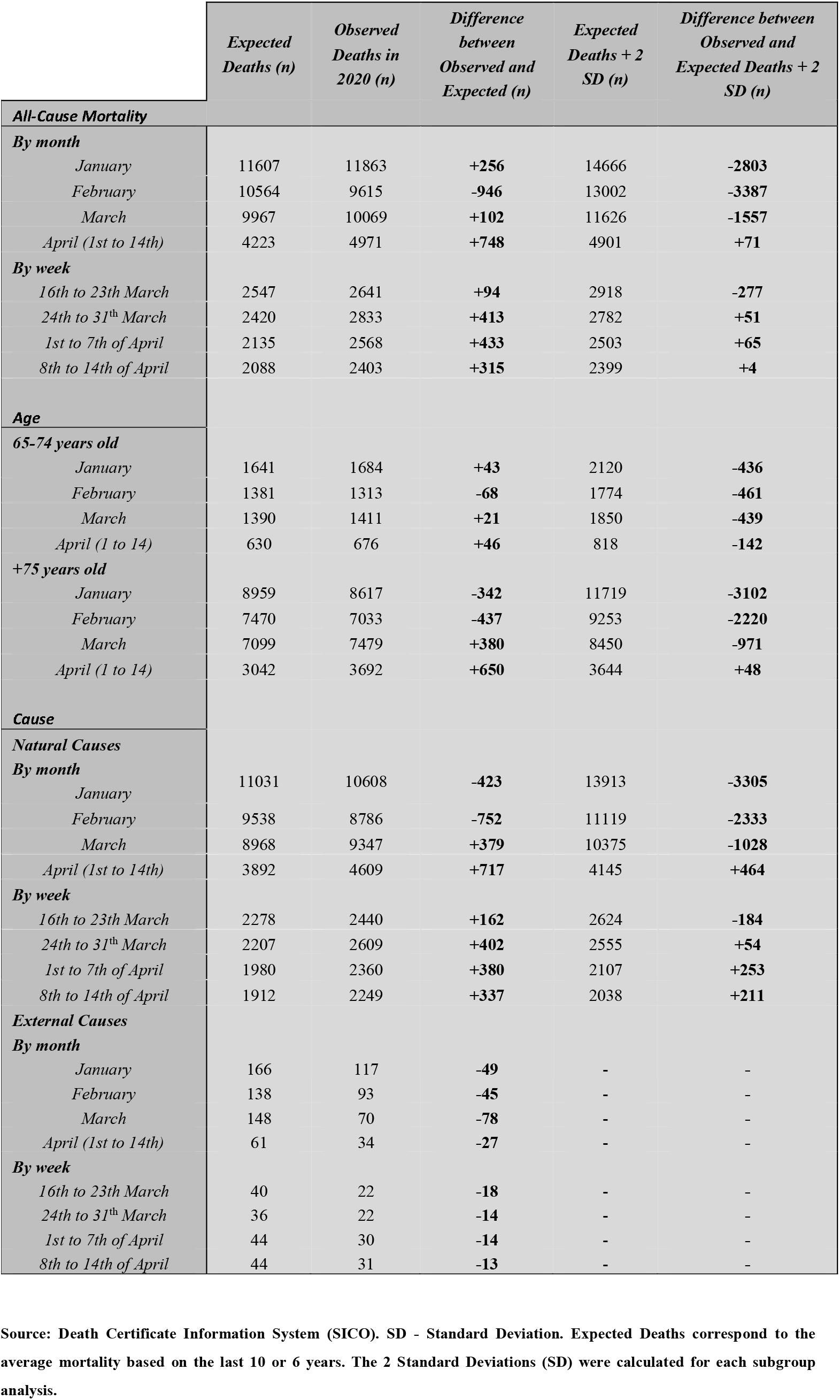
Overall description by all-cause, age and natural / external cause.

The estimated excess mortality has two components: 49% of excess deaths were caused by COVID-19 and another 51% were attributed to other causes. In the study period, 599 deaths were registered by COVID-19, so we estimate that the excess of non-COVID-19 Mortality (EMnC) was 615 deaths (1214 - 599), due to other causes natural causes not directly related to the pandemic.

## 3 Discussion

The findings of this study suggest an excess mortality during the first month of the COVID-19 pandemic in Portugal, with a gradual reduction of the trend from the second week in April.

The results seem robust as they are consistent using two different estimate methods: a) comparing observed and expected deaths, based on the calculation of the historical averages and 2SD of death records for the last 10 years, and b) using the ARIMA model, filling the historical average from the date were forecast would begin, we included historical data in ARIMA forecasting making the prediction and confidence levels closer to reality considering the actual year and the previous ones.

Our excess mortality estimates are conservative, since expected deaths are influenced by the previous years and the present year registered lower mortality than expected in February and early March. In addition, during the study period there was a significant reduction in the number of injury related deaths, namely those associated with traffic accidents, due to severe mobility restrictions imposed by the Authorities..^11^

The results are also in line with the estimates made by other excess mortality studies carried out in Portugal. Nogueira et al. estimated an EM between 2400 to 4000 deaths between March 1 and April 22.^12^ These differences can be attributed to different methods and time windows used.

These results are also in line with other and international studies, namely those of EuroMOMO.^13^ Another study reported significant excessive mortalities in New York City (+289%) and in countries like Spain (+66%), England and Wales (+33%), Netherlands (+33%) and Belgium (+25%).^4^ Difference may be explained, by different epidemic starting times, in different countries, the time at which each country adopted different confinement measures, and different compliance rates.^14^

Our study shows a decline in excess mortality two weeks after it implemented stringent confinement measures, with which the population complied. Other countries like England, Spain, Italy and the Netherlands have maintained very high levels of EM in weeks 13 and 14 of the respective epidemics.^3^

Our study suggests a concentration of excess mortality among those aged 75+. EuroMOMO also registers excess mortality for age groups between 15 and 64 years old and above 65. The difference may be related to the high lethality of the COVID-19 virus for patients aged 80+ (12.4%)^15^ and with the vulnerability of this population to changes in supply and demand for health care itself.

This study showed that 51% of the registered excess mortality was due to other natural causes, not classified as COVID-19 (EMnC). This may be explained by: a) some patients may have died with COVID-19 died at home, or at long-term care institution, without having had a COVID-19 test; other patients, with serious conditions, may not have sought hospital care, for fear of being infected, or did seek hospital care but did not get the full attention needed, because manpower and equipment were dedicated to the care of COVID-19 patients.

Under ascertainment may have been a problem during the first period of the study period and a study from the Imperial College of London about under-ascertainment estimated Portugal to report 36% of estimated number of infections^16^. This became less of a problem by April, when Portugal adopted a generous testing strategy, including tests in long term care institutions and even at the morgue. Finally, Portugal has an inclusive definition for classifying deaths as due to COVID-19, accepting the diagnosis in the absence of a confirmation test.^17^

A study by Rui Santana and collaborators (2020) supports the hypothesis that patients reduced emergency visits, since it showed that, in March 2020, there was a 48% drop in the demand for emergency services and a reduction of more than 144,000 emergency episodes, for patients with a higher priority of care (yellow and red bracelets), when compared to previous years.^18^

Some international studies also reported EM by non COVID-19 in different countries and found it varying from 20% to 60%, in Spain, Belgium France, England and Wales, Italy, the Netherlands, Switzerland and Sweden, which is in line with the results presented in this study.^19, 20^

Our study shows EM due to natural causes were superior to total EM. This difference partly explained by a significant reduction in the number of injury related deaths, namely those associated with traffic accidents, due to severe mobility restrictions imposed by the Authorities..^11^

Despite heterogeneous methods and findings of different studies, our results add to the body of evidence created around COVID-19 excess mortality.

## Conclusion

Between March 16 and April 14, 2020, there was an excess of 1 255 deaths over the expected, based on average daily mortality during the previous 10 years, an excess mortality of 13.7%. The number of deaths observed exceeded the defined threshold of Relevant Excess Mortality in 14 days of the study period, totaling 242 REM.

Almost all excess mortality (EM) was detected among people aged 75 years or more. Between March 1 and April 14, 2020, there was an EM of 1 030 deaths in people aged 75 +, compared to the daily averages of deaths in this age group, in the last 6 years. For this age group, the number of deaths observed exceeded the defined threshold of Relevant Excess Mortality in 11 days of the study period, totaling 201 REM.

Forty nine percent of excess mortality were caused by COVID-19 and another 51% were attributed to other natural causes. Collateral excess mortality by non COVID-19 needs further investigation

One month after the first death recorded by COVID-19 in Portugal, an EM is observed for all-cause mortality, mainly in the age groups above 75 years of age. Portugal registers less EM than many other countries. This may be due to the Authorities having decreed stringent confinement measures early and the high levels of compliance of the population.

As we enter a phase of deconfinement, it is necessary to maintain a high level of epidemiologic to be able to intervene early and with precision wherever there are signs of any excess morbidity or mortality by COVID-19.

## Data Availability

All data are available at SICO eVm website and DGS bulletins.

## Availability of data and materials

The datasets generated and/or analysed during the current study are available in the SICO eVm repository (https://evm.min-saude.pt/#shiny-tab-q_total) and Directorate-General of Health) daily Situation Reports (https://www.dgs.pt/informacao-e-comunicacao/historico-de-destaques.aspx).

## Key points

- An excess mortality of 1255 deaths were estimated one month after the first death classified by COVID-19, and it would probably be more if the government had not taken early action
- The age group where a significant increase in mortality was noted was above 75 years
- 51% of the EM was due to natural causes other than COVID-19

